# Association between tobacco exposure, blood pressure, and arterial stiffness in South African adults and children

**DOI:** 10.1101/2022.03.22.22272743

**Authors:** Betty Nembulu, Andrea Kolkenbeck-Ruh, Lebo F. Gafane-Matemane, Ruan Kruger, Yolandi Breet, Lisa J. Ware

## Abstract

**Introduction:** Sociodemographic factors, health status and health behaviour have all been associated with arterial stiffness. We examined the association between tobacco use or exposure and pulse wave velocity (PWV, a marker of arterial stiffness) in black South African adults and children against a background of other known risk factors.

**Methods:** Two datasets were used: African-PREDICT (A-P; n=587 apparently healthy black adult men and women, 20-30 years) and Birth-to-Twenty-Plus (Bt20; n=95 black adult women, 28-68 years and n=47 black children, 4-10 years). A cotinine value >10 ng/ml in urine (Bt20) or serum (A-P) was considered as tobacco exposed and carotid-femoral PWV was measured using the SphygmoCor XCEL device. Regression analysis included cotinine and other known risk factors.

**Results:** One third of adults (32%) and almost half of all children (45%) were tobacco exposed with the prevalence of elevated blood pressure (BP) approximately twice as high as their non-exposed counterparts (adults, p=0.014; children, p=0.017). Cotinine was the only variable that significantly associated positively with PWV in both adults and children in univariate analysis (p<0.05), but only MAP remained significant for adults in multivariate analysis (p=0.001).

**Conclusions:** In this sample, tobacco exposure was adversely associated with vascular health in adults and children. BP was higher in the tobacco exposed adults and children compared to their non-exposed counterparts. These findings suggest tobacco cessation programs for adults should include screening for blood pressure and consider the impact of tobacco exposure on children’s vascular health.

## Background

Sociodemographic factors, health status and health behaviour can all increase the risk of early vascular aging (EVA) and cardiovascular disease (CVD) (1). EVA is an independent predictor of target organ damage that indicate the deterioration in vascular structure and function (1). CVD is the leading cause of death globally, resulting in 17.9 million deaths annually (2). Identifying those at high risk for CVD remains a priority to prevent premature death. CVD results from vascular abnormalities as a result of complex interactions of different factors including genetics, environment, demographics and poor health behaviours such as tobacco use, which is a known preventable CVD risk factor (3–5). Tobacco use and exposure frequently coexist with other CVD risk factors, such that tobacco cessation efforts may present an opportunity to target multiple CVD risk reduction strategies if we know the associations with other risk factors. For example, healthy lifestyle choices such as cessation of tobacco use, regular physical activity and avoiding harmful use of alcohol have been shown to reduce CVD risk (2,6).

Tobacco is estimated to cause approximately 8 million deaths annually, with more than 7 million deaths attributed to direct tobacco use and approximately 1.2 million attributed to environmental tobacco exposure, also referred to as second hand smoke (7). In 2015, approximately 24.9% of the global population aged over 15 years were current users of tobacco (8). While tobacco use has declined globally, evidence has shown a relative increase in the African region (from 54 million tobacco users in 2010 to 57 million users in 2018) (8). Similar to the global prevalence, approximately 22% of the South African population aged 15 years and above are estimated to use tobacco products according to the 2016 South African demographic and health survey (9). Tobacco is frequently assessed through self-report, however, this method does not account for individuals who don’t smoke but are exposed to environmental tobacco smoke (10). Thus, cotinine, a major metabolite of nicotine, has become a reliable biomarker of both tobacco use and exposure (3,8).

Tobacco exposure can contribute to pathophysiological processes, such as endothelial dysfunction (11,12) that lead to EVA and arterial stiffness. There is a direct relationship between arterial stiffness and cardiovascular mortality (13). Vascular ageing, as commonly reflected by stiffening of large arteries, is a normal process that occurs with chronological ageing (12), due to increased elastin fragmentation and collagen accumulation leading to loss of elasticity and compliance of the arterial wall (12). However, increased arterial stiffness has been observed in South African children as early as 6 years of age (14). Arterial stiffness is determined by pulse wave velocity (PWV), a predictive value of aortic stiffness (13,15) whereby a PWV value ≥10 m/s in adults is a predictor of cardiovascular events (16). Evidence indicates that the risk for EVA starts earlier in life, however, limited evidence of vascular aging exist for children (1,17). As such, it is important to understand the impact of risk factors that are considered non-modifiable (age, sex, family history and socioeconomic status), as well as modifiable risk factors (tobacco exposure, excess adiposity, alcohol use, blood pressure and medication use) in the assessment of accelerated deterioration of the vasculature and to inform intervention strategies. Therefore, we examined the relationship between tobacco exposure, assessed by the biomarker cotinine, and PWV, a marker of arterial stiffness, against a background of other risk factors in black South African adults and children.

## Methods

This was a cross-sectional analysis of secondary data from two different cohort studies in South Africa. The first cohort, from Soweto, Gauteng province, was an intergenerational study on the transmission of vascular health in families from the Birth-to-Twenty (Bt20) cohort with data collected between August 2019 to March 2020. The second cohort was from Potchefstroom, North West province, the African Prospective study on the Early Detection and Identification of Cardiovascular disease and Hypertension (African-PREDICT) with data collected between 2013 and 2017. Inclusion criteria for Bt20 were: BT20 index females, with both a biological child aged between 4-10 years and a biological mother still alive, to be able to determine the original aim of this study i.e. the degree to which vascular health is transmitted through generations. Individuals with apparent physical, mental, or congenital abnormalities were excluded. For this analysis, only participants with complete vascular health measurements who gave consent (adults) or assent in all the children aged 7-10 years for the collection of urine samples and analysis of urinary cotinine were included. Inclusion criteria for African-PREDICT were: 20-30 years old; male or female; black or white ethnicity; and apparently healthy with a BP measurement <140/90 mmHg. Individuals with pre-existing cardiovascular disease were excluded. However, only black participants (female and male) with complete vascular health measures who gave consent for serum samples were included for this analysis. The total sample size for this analysis was 729 participants, inclusive of children (n=47; 4-10 years from Bt20) and adults (n= 682; 19-68 years), **(Figure 1)**.

**Figure 1.**
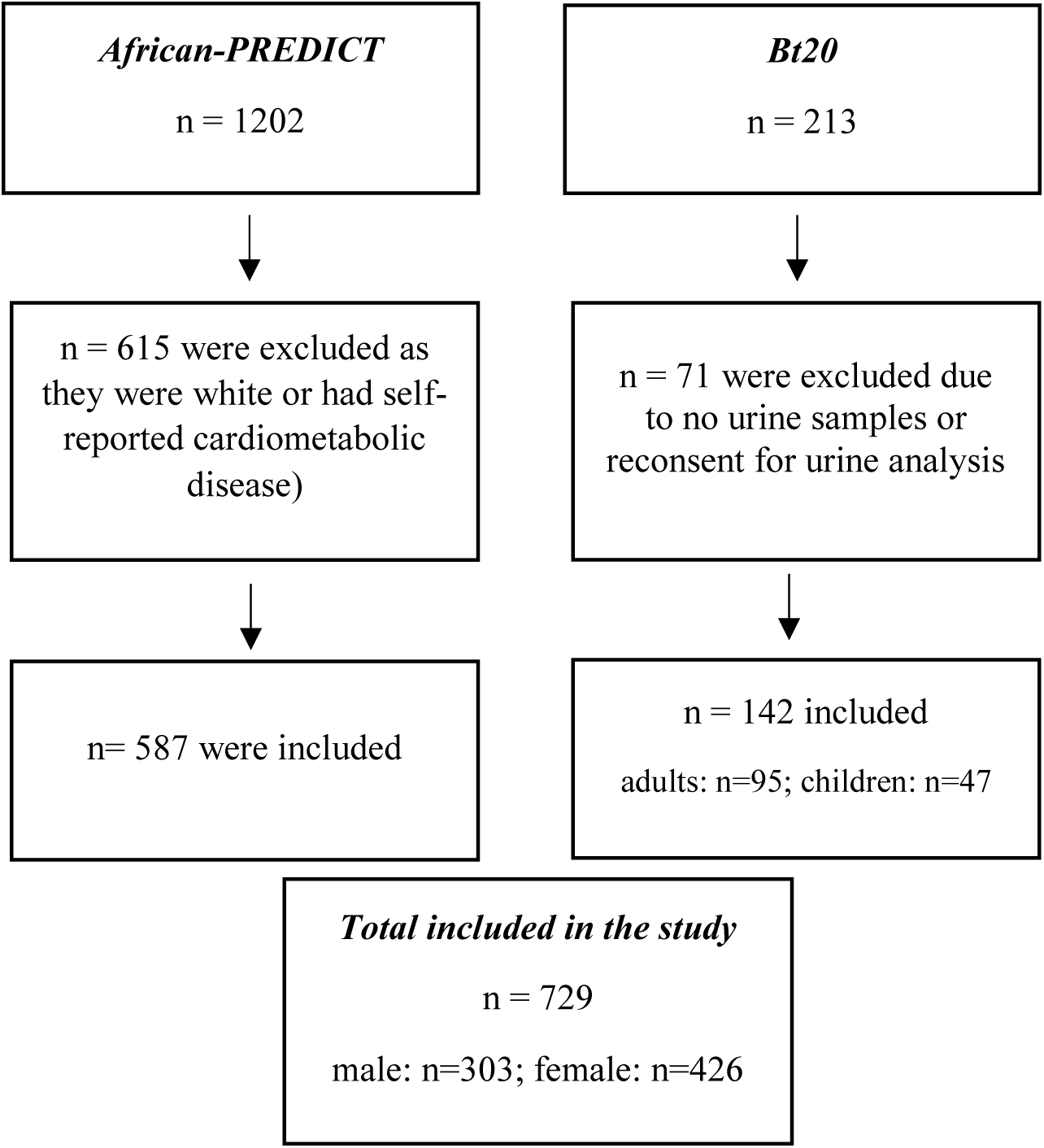
Study flow diagram

## Ethical consideration

Ethical approval was obtained from the Witwatersrand Human Research Ethics Committee (Medical) (M190263) and the North-West University Health Research Ethics Committee (NWU-HREC) of the Faculty of Health Sciences (NWU-00001-12-A1). Informed consent was obtained from both Bt20 and African-PREDICT adults and assent in all the Bt20 children aged 7-10 years.

## Measurements

### Health questionnaire

In both studies a general health and demographic questionnaire was used to obtain self-reported health data and demographics, including age and sex. Socio-economic data, family history and alcohol use were recorded through a health questionnaire, while tobacco use was assessed using the Global Adult Tobacco Survey for Bt20 only (18). Self-reported medical history (type 2 diabetes mellitus (T2DM), hypertension, high cholesterol, or previous heart disease) and current medication use, including anti-hypertensive, diabetic and high cholesterol medication, was assessed by questionnaire in both studies.

### Anthropometry

Height, weight and waist circumference, were measured in triplicate using the standardized World Health Organization (WHO) measurement protocol (19) and the International Society for the Advancement of Kinanthropometry protocol (20) for Bt20 and African-PREDICT studies respectively. Thereafter, waist to height ratio (WHtR) was calculated in both adults and children, where a WHtR 0.50-0.59 was classified as overweight and ≥ 0.60 as obese (21).

### Blood pressure measurements

Brachial blood pressure (BP) was measured using automated devices (Bt20 – Omron MIT5 for adults; Omron HBP-1300 for children (22); Omron Healthcare, Kyoto, Japan; African-PREDICT BP - Dinamap® Procare BP monitor, Milwaukee, US). Three measures were taken and the second and third measurement on the right arm averaged following the International Society of Hypertension measurement guidelines (23). Participants were seated and asked to rest for at least 5-minutes prior to measurement in a quiet room with the monitor facing away from the participant and a 2-minute rest interval between the measurements. Elevated BP was defined as ≥90th percentile for children (24) and SBP ≥130 mmHg and/or DBP ≥85 mmHg in adults (23). The right brachial mean arterial pressure (MAP) was calculated using the following equation: MAP = DBP + 1/3(SBP – DBP) (25).

### Pulse wave velocity (PWV) measurements

All participants were asked to complete an overnight fast and to avoid tobacco use prior to measurements. Participants rested in a supine position for at least 10-minutes prior to PWV measurements. We used the SphygmoCor XCEL device (AtCor Medical, Sydney, New South Wales, Australia) to measure central BP and PWV. The distance between the carotid and femoral arteries was measured with a tape measure using the direct distance method, as recommended by an expert consensus on the measurement of aortic stiffness in 2012 (16), and using 80% of the distance from the carotid pulse to the femoral pulse. Peripheral BP was obtained by partially inflating a size appropriate cuff over the right brachial artery approximately midway between the shoulder and the elbow. This was needed as an input variable along with height for PWV measurement. Thereafter, to assess carotid-femoral PWV, following input of the participant’s peripheral BP, height, sex and date of birth, carotid pulse waves were measured by applanation tonometry (high-fidelity micromanometer; Millar Instruments) while a partially inflated cuff obtained femoral pulse waves over the femoral artery at the leg midway between the hip and the knee, simultaneously capturing carotid and femoral waveforms over a pre-set time of 10-seconds.

### Cotinine samples

For Bt20, all participants were asked to produce spot urine on the same day the PWV measurements were performed. Three 1.5ml aliquots were taken for each participant and stored at -80°C prior to biochemical analysis. For African-PREDICT, blood samples were drawn from the antebrachial vein, centrifuged in an onsite laboratory to obtain serum aliquots and stored at -80°C until analysis (26). Urine and serum cotinine were analysed in different laboratories but using the same chemiluminescence method on the IMMULITE system (Siemens, Erlangen, Germany). For both urine and serum, samples above the analytical cotinine limit of detection (LOD; 10 ng/ml) were classified as tobacco exposed (either through use or environmental exposure) as previously shown to be a valid threshold in this population (27). When used as a continuous variable in the regression analysis, to avoid excluding concentrations below the LOD (<10 ng/ml), these were substituted by dividing the LOD by two (10/2=5 ng/ml), a method shown to reduce bias (28) and values were log transformed.

## Statistical analysis

Normality of data was tested using visual inspection of histograms and the Shapiro-Wilks test, with continuous variables reported as mean ± standard deviation (SD) or median and interquartile range (IQR). Differences between tobacco-exposed and non-exposed adults and children were tested using Chi-square tests for categorical data, and paired T-tests (parametric) or Mann-Whitney U (nonparametric) for continuous data. Analysis of covariance (ANCOVA) was then used to adjust PWV for MAP to assess differences in study participants by tobacco exposure category. Linear regression analyses (univariate and multivariate) were used to explore the relationship between PWV, cotinine, and other known risk factors (MAP, antihypertensive medication use, contraception use, self-reported alcohol use, and WHtR, age, sex, family history, and education level) in children and adults, and for adults with urine and serum cotinine separately. SPSS version 27.0 (IBM Corp, Armonk, NY) was used for all statistical analyses.

## Results

### Study participants characteristics

**Table 1** summarizes participant characteristics by tobacco exposure group. In adults, 31% were classified as tobacco exposed. Overall, the tobacco exposed adult group were slightly older (*p*=0.018), more frequently male (*p*<0.001), with higher reported alcohol use (*p*<0.001) and increased prevalence of elevated blood pressure (*p*=0.014) and antihypertensive medication use (*p*=0.017). Prevalence of hypertension in the adults was low (<10%) as African-PREDICT excluded adults with elevated blood pressure at baseline. Tobacco exposed adults had significantly higher PWV when adjusted for MAP (p=<0.001).

**Table 1.** Characteristics of adults and children by tobacco exposure categories

| Characteristics | Adults (n=660) |  | Children (n=47) |  |
| --- | --- | --- | --- | --- |
|  | Tobacco exposed<sup>#</sup><br>(n=209) | Tobacco not<br>exposed<sup>#</sup> (n=451) | Tobacco exposed<sup>#</sup><br>(n=21) | Tobacco not<br>exposed<sup>#</sup> (n=26) |
| ***Socio-demographics*** |  |  |  |  |
| Age (years) | 26.0 (6.0) | 25.0 (6.0)* | 6.0 (3.0) | 7.5 (2.3) |
| Females, n (%) | 98 (46.9) | 289 (64.1)*** | 12 (57.1) | 16 (61.5) |
| Males, n (%) | 111 (53.1) | 162 (35.9)*** | 9 (42.9) | 10 (38.5) |
| ***Socioeconomic status*** |  |  |  |  |
| Education level (years) | 8.0 (2.0) | 8.0 (2.0) | - | - |
| ***Anthropometry*** |  |  |  |  |
| Height (cm) | 165.1±8.3 | 163.0±8.4 | 120.8±11.1 | 126.3±11.6 |
| Weight (cm) | 64.3 (18.5) | 65.7 (18.0) | 22.5 (9.6) | 23.8 (13.4) |
| Waist circumference (cm) | 77.0 (20.0) | 77.6 (15.0) | 54.7 (8.7) | 55.3 (14.7) |
| WHtR | 0.5 (0.1) | 0.5 (0.1) | 0.5 (0.1) | 0.5 (0.1) |
| WHtR class |  |  |  |  |
| Obese (WHtR ≥0.60), n (%) | 35 (16.7) | 51 (11.3) | 0 (0) | 1 (3.8) |
| Overweight (WHtR 0.50-0.59), n (%) | 46 (22.0) | 118 (26.2) | 5 (23.8) | 5 (19.2) |
| Normal (WHtR < 0.50), n (%) | 81 (38.8) | 172 (38.1) | 15 (71.4) | 19 (73.1) |
| ***Medical history*** |  |  |  |  |
| Self-report Hypertensive, n (%) | 15 (7.2) | 15 (3.3)* | - | - |
| Self-report high cholesterol, n (%) | 5 (2.4) | 7 (1.6) | - | - |
| Self-report diabetes mellitus, n (%) | 1 (0.5) | 2 (0.4) | - | - |
| Family history, n (%) | 114 (54.5) | 239 (53.0) | - | - |
| Antihypertensive medication use, n (%) | 13 (6.2) | 13 (2.9)* | - | - |
| Contraceptive use, n (%) <sup>#</sup> | 7 (3.3) | 48 (10.6) | - | - |
| ***Health behaviours*** |  |  |  |  |
| Cotinine, ng/ml | 170.0 (254.0) | <10*** | 18.0 (46.8) | <10*** |
| Current alcohol consumption, n (%) | 149 (71.3) | 205 (45.5)*** | - | - |
| Current tobacco use, n (%) | 119 (56.9) | 26 (5.8)*** | - | - |
| ***Right seated office blood pressure*** |  |  |  |  |
| Systolic BP (mmHg) | 117 (17) | 116 (18) | 108 (16) | 102 (10) |
| Diastolic BP (mmHg) | 77 (14) | 79 (12) | 71 (11) | 69 (10) |
| Pulse pressure (mmHg) | 39 (11) | 40 (11) | 35 (11) | 43 (24) |
| Right brachial MAP (mmHg) | 92 (13) | 78 (12) | 83 (8) | 81 (11) |
| BP status |  |  |  |  |
| Elevated BP, n (%) <sup>‡</sup> | 18 (8.6) | 18 (4.0)* | 18 (85.7) | 12 (46.2)* |
| Normotensive BP, n (%) | 188 (90.0) | 431 (95.6)* | 3 (14.3) | 11 (42.3)* |
| ***SphygmoCor data*** |  |  |  |  |
| Central systolic pressure (mmHg) | 111 (17) | 111 (13) | 92 (9) | 93 (12) |
| Central diastolic pressure (mmHg) | 76 (10) | 77 (11) | 63 (7) | 65 (5) |
| Central pulse pressure (mmHg) | 34 (9) | 33 (7) | 29 (5) | 26 (8) |
| PWV (m/s) | 6.7 (1.4) | 6.3 (1.1)*** | 4.0 (0.5) | 4.3 (0.7) |
| PWV adjusted for MAP | 6.7 (1.4) | 6.3 (1.1)*** | 4.0 (0.5) | 4.3 (0.7)* |

Data presented as median and interquartile range or mean ± standard deviation. T-test was used on normally distributed variables, Mann Whitney u-test for non-normally distributed variables and Chi-squared for categorical variables. <sup>#</sup> Contraceptive use includes both hormone replacement therapy and contraceptives. <sup>‡</sup>Elevated BP: ≥90th percentile for children(24) and SBP ≥ 130 mmHg and/or DBP ≥85 mmHg in adults(23). BP, blood pressure; MAP, mean arterial pressure; PWV, pulse wave velocity; WHtR, waist to height ratio. \*p<0.05; \*\*p<0.01; \*\*\*p<0.001

In children, 45% were classified as tobacco exposed. The tobacco exposed children had lower MAP adjusted PWV (*p*=0.046), but increased prevalence of elevated blood pressure (*p*=0.017) compared to the tobacco not exposed group.

### Linear regression analysis

Results of the linear regression analysis in children and adults (**Table 2)** showed that in children, only cotinine was significant for predicting PWV in the univariate analysis but not in multivariate analysis. In adults, cotinine together with anti-hypertension medication, contraception use, WHtR, age, sex and education level were significant for predicting PWV in the univariate analysis but in the multivariate analysis only MAP was significant (*p*<0.001). In the multivariate analysis for adults, for each one unit (mmHg) increase in MAP, there was an increase in PWV of 0.040 m/s.

**Table 2.**
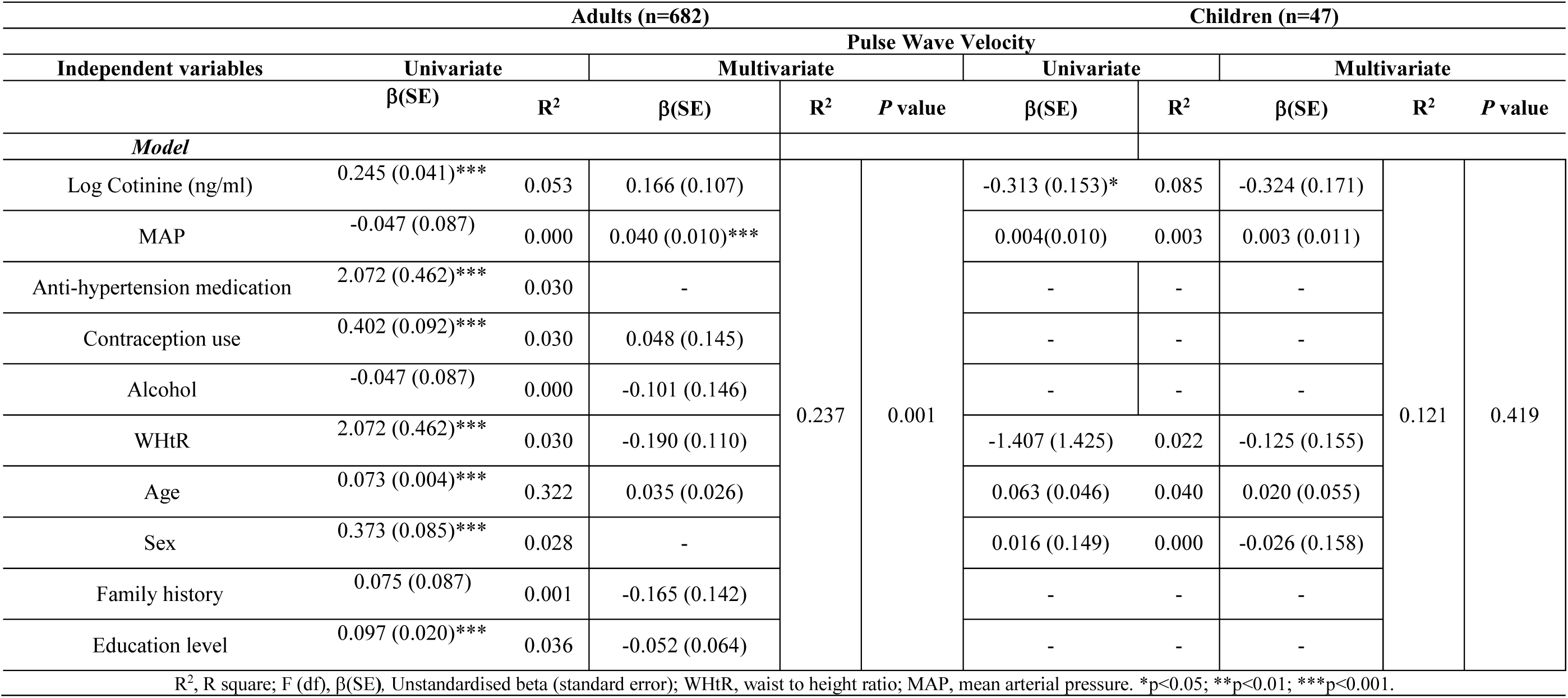
Linear regression analysis to evaluate the relationship between tobacco exposure and pulse wave velocity taking into consideration current health status, current modifiable health behaviours and current non-modifiable risk factors in adults and children

When the results for urine and serum cotinine in adults were analysed separately (**Table 3)**, the results showed that serum cotinine was significant (*p*<0.001) in the univariate analysis in the prediction of PWV. While MAP was significant (*p*<0.001) in the multivariate analysis, with the model explaining 24% of the variance in PWV. In adults with urine cotinine, no significant association was found between cotinine and PWV in either the univariate (*p*=0.714) or multivariate (*p*=0.930) analysis, though the sample size for this group was much smaller (n=95 with urine cotinine; n=587 with serum cotinine).

**Table 3.**
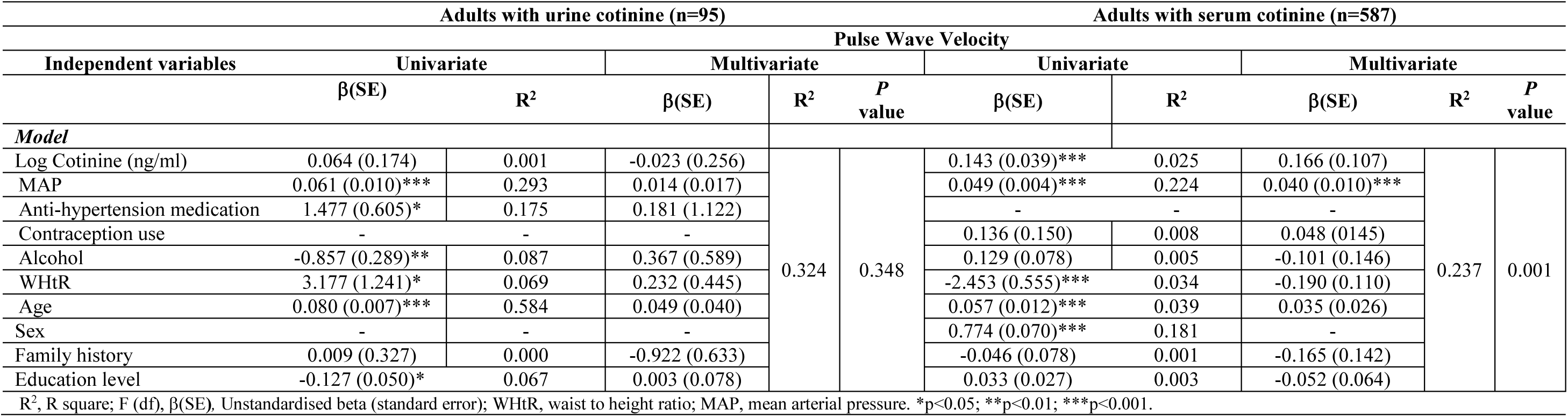
Linear regression analysis to evaluate the relationship between tobacco exposure and pulse wave velocity taking into consideration current health status, current modifiable health behaviours and current non-modifiable risk factors in adults

## Discussion

Our aim was to evaluate the number of adults and children who used or were exposed to tobacco as indicated by the biomarker cotinine and the association between cotinine and arterial stiffness. We found that one third of adults had cotinine detected in either their urine or serum and almost half of children had cotinine detectable in their urine. In both adults and children classified as being exposed to tobacco, the frequency of elevated BP was doubled, while PWV was higher in adults only. Tobacco exposure was additionally a significant predictor of PWV in univariate analysis in both adults and children, while in multivariate analysis blood pressure (MAP) predicted arterial stiffness in adults only.

When considering the results from adults with either urinary or serum cotinine, prediction of PWV was stronger in the serum group. This may indicate that urine cotinine may not be as good to detect associations with markers of vascular health as serum cotinine. although urinary cotinine has been established previously as a good detector of tobacco exposure in a South African adult population (29). This may be influenced by the metabolic enzymes in the liver responsible for oxidation of cotinine (such as CYP2A6 and a cytoplasmic aldehyde oxidase) (30). These enzymes can be influenced by age, sex, hormones, medication use and kidney disease (30).

Our results indicated that MAP was significant in the prediction of PWV in adults. These findings support previous research showing the association between blood pressure and tobacco exposure in the prediction of PWV in adults (31). One South African study found that, when compared to Caucasian smokers, African smokers showed higher arterial stiffness, indicating greater impact of tobacco use on cardiovascular dysfunction (32). The association between tobacco exposure and CVD has been well established in developed countries but there is still limited data in low- and middle-income countries (LMICs) like South Africa (32). However, more than 80% of the 1.3 billion tobacco users worldwide are based in LMICs (7).

A review in 2016 suggested that nearly 100,000 children transition to regular smoking daily, with 14,000–15,000 children per day in high income countries (HICs) and 68,000–84,000 in LMICs (33). To delay the onset of poor vascular health and deterioration such as arterial stiffness, health behaviours and exposures need to be changed early in life (5,31), indicating interventions are required to maintain ideal cardiovascular health. However, multi-pronged approaches are needed involving public health science, policy, and clinical practice targeted at the reduction of tobacco use; promoting cardiovascular health; and preventing tobacco-attributable deaths from CVD (33).

While the WHO Framework Convention on Tobacco Control (WHO FCTC) implemented as the Global Strategy to Accelerate Tobacco Control has been progressive over the years (34), challenges have been experienced in LMICs (33). These have been cited as, slow integration into national law, insufficient funds for implementation and tobacco industry interference with policy-making (34).

South Africa passed comprehensive national legislations on tobacco controls, which are compliant with the WHO FCTC (35). The legislations cover increasing tobacco prices, the use of health warnings on packaging, smoke free policies and anti-smoking messaging via mass media marketing, which have been effective in the reduction of tobacco use (7,33,36). Despite the tobacco control regulations put in place, there is still a need for programs to help smokers who want to quit, and training programs to educate healthcare providers to help active smokers break their nicotine addiction.

## Limitations

A limitation of the study was the small sample of children (n= 47) as compared to adults (n= 682), with lower absolute cotinine values in children that may influence our findings related to arterial stiffness. In addition, the sample of adults with urinary cotinine was smaller than the sample with serum cotinine. In children only urine cotinine was available. One other limitation of this study is that urine and serum cotinine were not analysed in the same laboratory, although the same method and equipment were used. Furthermore, overall prevalence of hypertension in adults was low (<10%) as African-PREDICT aimed to evaluate change in cardiovascular health over time excluding adults with elevated blood pressure at baseline. In a representative sample of adults where hypertension prevalence would be higher, the impact of tobacco exposure on hypertension and vascular health may be greater. The strengths of this study were the large overall sample size (n=729), with measurements taken in controlled environments following the same standard procedures and analytical methods.

## Conclusion

In this study, one third of black South African adults and almost half of the children aged 4-10 years were found to have cotinine in either their serum or urine, indicating use of tobacco and/or exposure to tobacco users. The prevalence of elevated blood pressure was approximately twice as high in those adults and children exposed to tobacco. In adults, blood pressure appeared to associate most closely with arterial stiffness. Our findings indicate that tobacco cessation strategies in South Africa should consider other cardiovascular risk factors, especially at younger ages and include blood pressure screening of those using tobacco and those exposed to tobacco use within their households.

## Data Availability

Data is available on request from Professor Shane Norris.

## Acknowledgements

Funding support for the Birth to Twenty Plus Cohort has been received from the South African Medical Research Council, Wellcome Trust, and DSI-NRF Centre of Excellence in Human Development at the University of the Witwatersrand, Johannesburg, South Africa. Non-communicable disease research unit (NCDR) financial contribution is hereby acknowledged.

Funding support for African-PREDICT has been received from the South African Medical Research Council (SAMRC) with funds from national treasury; the South African Research Chairs Initiative (SARChI) of the Department of Science and Technology and National Research Foundation (NRF) of South Africa (GUN 86895); the SAMRC with funds received from the South African National Department of Health; GlaxoSmithKline R&D (Africa non-communicable disease open lab grant); the UK Medical Research Council and with funds from the UK Government’s Newton Fund; corporate social investment grants from Pfizer (South Africa), Boehringer Ingelheim (South Africa), Novartis (South Africa), the MediClinic Hospital Group (South Africa); and contributions from Roche Diagnostics (South Africa).

## Notes

**Declaration of interests** No conflicts of interest.

**Funding** This work was supported by a Wellcome Trust grant awarded to Dr. Lisa Ware [214082/Z/18/Z].

### Competing Interest Statement

The authors have declared no competing interest.

### Funding Statement

This work was supported by a Wellcome Trust grant [214082/Z/18/Z].

### Author Declarations

Ethical approval was obtained from the Witwatersrand Human Research Ethics Committee (Medical) (M190263) and the North-West University Health Research Ethics Committee (NWU-HREC) of the Faculty of Health Sciences (NWU-00001-12-A1).

